# The effect of exogenous ketones on signs and symptoms of schizophrenia spectrum and bipolar disorders: study protocol for a triple-blind, randomized, controlled cross-over pilot study

**DOI:** 10.1101/2025.03.02.25323118

**Authors:** Daphne AM Dielemans, Yagmur Yurtkap, Marieke van der Pluijm, Maarten R Soeters, Bob Oranje, Dirk JA Smit, Tim Ziermans, Mirjam J van Tricht, Sriram Muthukumar, Shalini Prasad, Romée L van der Mieden van Opmeer, Eline Dekeyster, Astrid M Kamperman, Jason RB Dyck, Bram-Sieben Rosema, Rocco Hoekstra, Ralph W Kupka, Lieuwe de Haan, Nico JM van Beveren, Karin Huizer

**Affiliations:** Parnassia Psychiatric Institute, The Hague; Amsterdam University Medical Center, dept. of Psychiatry; Amsterdam University Medical Center, dept. of Endocrinology and Metabolism; Academic Hospital Glostrup, Center for Neuropsychiatric Schizophrenia Research; University of Amsterdam, D utch Autism & ADHD Research Center; GGZ Centraal, Laren, dept. of Neuropsychiatry; Enlisense LLC, Allen, Texas; University of Texas at Dallas, dept. of Bioengineering; Leiden University, dept. of Cognitive Psychology; Erasmus Medical Center, Rotterdam, Epidemiological and Social Psychiatric Research Institute, dept. of Psychiatry; University of Alberta, Edmonton, Cardiovascular Research Centre, dept. of Pediatrics; University Medical Center Groningen, dept. of Psychiatry; Erasmus Medical Center, Rotterdam, dept. of Pathology

**Keywords:** exogenous ketones, ketone ester, schizophrenia, bipolar disorder, ketosis, treatment, prepulse inhibition

## Abstract

**Introduction:** Inflammation, oxidative stress, and bioenergetic dysfunction are proposed underlying mechanisms of schizophrenia spectrum disorders (SSD) and bipolar disorders (BD), contributing to the largely untreated cognitive and negative symptoms in these conditions. Ketone bodies may offer a therapeutic option for these symptoms through their positive effects on the aforementioned mechanisms. Exogenous ketones like ketone esters (KE) provide a means to quickly induce ketosis without dietary restrictions, though their effects on SSD and BD have not yet been investigated. Here, we describe the study protocol of an ongoing triple-blind, randomized controlled crossover trial on the effects of a single ingestion of KE on signs and symptoms of SSD and BD.

**Methods:** A total of 24 patients (12 SSD, 12 BD) receiving inpatient care at Amsterdam UMC will be included in the study. Patients will ingest a single dose of KE ((R)-3-hydroxybutyl-(R)-3-hydroxybutyrate deltaG® ketones - dGK) and an isocaloric carbohydrate control with a washout period of three days between drinks. The primary outcome is the change in pre-pulse inhibition of the startle reflex (PPI) induced by dGK ingestion compared to control. Secondary outcomes include resting-state EEG, P3B amplitude, cognitive performance, and metabolic, immune, oxidative stress and circadian rhythm parameters. Feasibility and potential side effects will also be assessed.

**Discussion:** Our current study offers valuable preliminary data on the effects of KE in SSD and BD patients. It can provide the foundation for future research into the therapeutic potential of KE in alleviating symptoms and improving functional outcomes in these disorders.

**Trial registration:** www.ClinicalTrials.gov, ID: NCT06426134.

## 1. Introduction

Schizophrenia spectrum disorders (SSD) and bipolar disorders (BD) are severe mental disorders associated with a high disease burden and increased mortality [1]. SSD is characterized by positive symptoms (e.g., hallucinations, delusions), negative symptoms (e.g., lack of motivation, social withdrawal), and cognitive symptoms (e.g., difficulties with executive functions and working memory). BD involves (hypo)manic episodes often alternating with depressive episodes. Antipsychotic medication, which mainly inhibit the dopamine D_2_ receptor, are an important form of treatment for both disorders. The effectiveness of dopamine receptor antagonists in alleviating positive symptoms of SSD has been pivotal in the development of the dopamine hypothesis [2].

While antipsychotics are effective in reducing positive and manic symptoms, they fail to address cognitive [3-8] or negative [9, 10] symptoms, which significantly contribute to the chronic disease burden of SSD or BD. Additionally, antipsychotics can generate secondary negative symptoms, such as affect flattening and lack of motivation, as well as cognitive dysfunction at higher doses, thereby exacerbating the burden of negative and cognitive symptoms [11, 12].

Besides improving cognition and negative symptoms, another unmet treatment need concerns modification of etiopathophysiological mechanisms. Mechanisms such as glutamatergic dysregulation, oxidative stress, and inflammation are thought to play a central role in SSD and BD [13-15]. The glutamate hypothesis posits that disruptions in glutamatergic signaling, particularly involving N-methyl-D-aspartate (NMDA) receptors, contribute to the symptoms of SSD and BD [14, 16]. Oxidative stress refers to an imbalance between the production and neutralization of reactive oxygen species (ROS). Excessive ROS induce oxidative damage to cellular macromolecules, including lipids, proteins and DNA, resulting in impaired neuronal function and synaptic dysfunction [13]. Oxidative stress and inflammation are interconnected mechanisms often described together in the context of SSD and BD. Recent evidence suggests that disruptions in glucose metabolism may underlie these disorders [17-19]. According to the bioenergetic dysfunction hypothesis, the capacity of the brain to efficiently utilize glucose for ATP production may be impaired. Reduced glycolytic enzyme activity and mitochondrial dysfunction have been suggested as underlying mechanisms [19]. This disrupted energy homeostasis is thought to impair neuronal communication, contributing to cognitive deficits and negative symptoms [18, 19].

Nutritional ketosis (NK)—an increase in blood ketone levels through nutritional interventions—may offer clinical benefits for SSD or BD by improving bioenergetic deficits [20, 21]. NK can be induced through fasting, consumption of medium-chain triglycerides (MCT), ingestion of exogenous ketones, or adherence to a ketogenic diet (KD), which is characterized by high fat and low carbohydrate intake [22-24]. In these situations, ketone bodies (primarily β-hydroxybutyrate (BHB) and acetoacetate (AcAc)), are produced via the β-oxidation of fatty acids in the liver (ketogenesis) and released into the blood stream. It commonly takes several days to a few weeks to fully adjust to a KD. Contrarily, when KE is ingested, ketone bodies become immediately available as an energy source upon resorption. The brain can effectively use ketone bodies as an alternative energy source: ketone bodies are efficiently transported across the blood-brain barrier [25, 26] and metabolized in neural mitochondria for ATP production [21]. Furthermore, KE intake in non-fasting mice resulted in increased brain ketone uptake and reduced glucose metabolism [27], suggesting that ketones are preferentially utilized when available. NK is also thought to improve neuronal energy metabolism by stimulating mitochondrial biogenesis, which enhances mitochondrial function and ATP production [22, 23]. By supplying ketone bodies, the brain may therefore bypass energy metabolism defects [28] observed in SSD and BD patients.

The effects of NK on the brain extend beyond providing an alternative energy source to glucose for neuronal mitochondria. Ketone bodies can mitigate oxidative stress and inflammation, by increasing the NAD+/NADH ratio in mitochondria, thus decreasing ROS production and enhancing ATP synthesis [29]. Additionally, the antioxidant properties of ketone bodies may be effective in reducing neuronal cell death [30]. Given the close relationship between oxidative stress and inflammation, the KD may indirectly modulate the immune responses by reducing oxidative stress. However, ketone bodies are can directly target the inflammatory system through various pathways [23, 31]. Indeed, the KD has demonstrated neuroprotective effects by inhibiting inflammation induced by macrophages and microglia in the central nervous system [32]. Finally, ketone bodies also suppress transcription of pro-inflammatory cytokines involved in SSD and BD (IL6, TNFA, IL1B)-[20]. Furthermore, ketone bodies influence glutamate metabolism by increasing glutathione levels [22] and can reduce glutamate-induced excitotoxicity[33, 34]. In summary, previous studies have highlighted multiple mechanisms through which NK may exert its effects, including its impact on mitochondrial energy production, oxidative stress, inflammation, and neurotransmitter levels. These mechanisms overlap with key pathophysiological processes involved in SSD and BD, underscoring the potential of NK as a therapeutic approach for these disorders.

In clinical research, a KD has demonstrated potential for improving cognitive function [35-37], modulating the immune system [38, 39], and addressing metabolic syndrome [40, 41] in various non-psychiatric contexts. Some findings indicate improved sleep quality following a KD, possibly by influencing the circadian rhythm through ketone bodies [34]. While the KD is an evidence-based treatment modality for treatment-resistant epilepsy [42, 43], research into its efficacy in psychiatric diseases like SSD or BD is still in its early stages as reflected by published results deriving from small and uncontrolled trials [44-49]. While a blinded RCT on the effects of the KD in SSD is currently ongoing [50], a knowledge void remains regarding the potential positive effects of NK on cognitive and negative symptoms in SSD/BD. Data from SSD animal studies support positive effects of ketones on deficits in sensorimotor gating (a process essential for adequate cognitive functioning), as measured by pre-pulse inhibition of the startle reflex (PPI) [51, 52]. A single dose of exogenous ketones could reverse PPI deficits in this animal model [53]. Since PPI is often disrupted in SSD and BD [54, 55], this is a promising finding.

While a KD may be a realistic option to achieve ketosis for some, its strict carbohydrate restriction is generally considered a major hurdle often leading to low or moderate adherence [56, 57]. When acute psychiatric episodes or negative symptoms like loss of motivation are present, a KD is usually not a valid option. On the other hand, the ingestion of KE can induce acute and potent ketosis without the need for a challenging diet [27, 58]. The KE (R)-3-hydroxybutyl (R)-3-hydroxybutyrate (deltaG® Ketones, dGK) has been available as a food product for consumers for several years. Research indicates its potential efficacy in improving outcomes in conditions such as Alzheimer’s disease, Parkinson’s disease, and type 2 diabetes, as well as its application in enhancing athletic performance [59-61]. To the best of our knowledge, no studies to date have evaluated the possible efficacy of KE supplementation in SSD or BD patients.

Here, we present a research protocol for a pilot study currently conducted at Amsterdam UMC, the Netherlands, which investigates the effects of KE ingestion in patients with SSD or BD. Our current study is part of a broader research initiative aimed at identifying novel therapeutic strategies for addressing negative and cognitive symptoms in SSD and BD.

### 2. Methods

### 2.1 Ethics Statement

The study has been approved by the Amsterdam UMC Institutional Review Board (IRB/METC AUMC) under the number 2024.0100, in accordance with the Medical Research Involving Human Subjects Act (WMO) which is based on the Nuremberg Code, the Declaration of Helsinki (version October 2013) and the ICH Good Clinical Practice guideline (ICH-GCP).

The study is registered at Clinicaltrials.gov with the following ID: NCT06426134. The study is conducted in accordance with the local legislation and institutional requirements. The participants provide their written informed consent to participate in this study.

### 2.2 Aims and objectives

The aim of this pilot study is to investigate the effects of a single dose of dGK on signs and symptoms of SSD or BD and to explore possible mechanisms of action. The primary outcome measure is the change in pre-pulse inhibition (PPI) following dGK ingestion compared to an isocaloric carbohydrate control drink in the same patient with SSD or BD. Secondary outcomes include resting-state EEG, P3B amplitude, cognitive performance, and patient-reported measures on mood, energy levels, and focus. Additionally, immune, oxidative stress, metabolic and circadian rhythm parameters are assessed. We will also evaluate the feasibility of dGK ingestion by examining its palatability and potential side effects in this patient group. This pilot study is intended as a lead-up for a future larger trial with a longer KE administration and follow-up.

### 2.3 Trial design and setting

This pilot study is designed as a triple-blind, randomized controlled cross-over trial. An isocaloric carbohydrate control is used to account for the potential effects of caloric intake on outcome measures and to control for any placebo effects. Both study drinks are generally considered to have an unpalatable taste and texture, aiding in blinding. The total study duration is five days, consisting of baseline measurements, two test days, and several continuous assessments. The KE or control drink is administered on day 2 of the study. After a 72-hour washout period, participants switch to the alternate condition on day 5. An overview of the scheduled measurements is provided in Figure 1, with a more comprehensive description presented in a later section.

**Figure 1.**
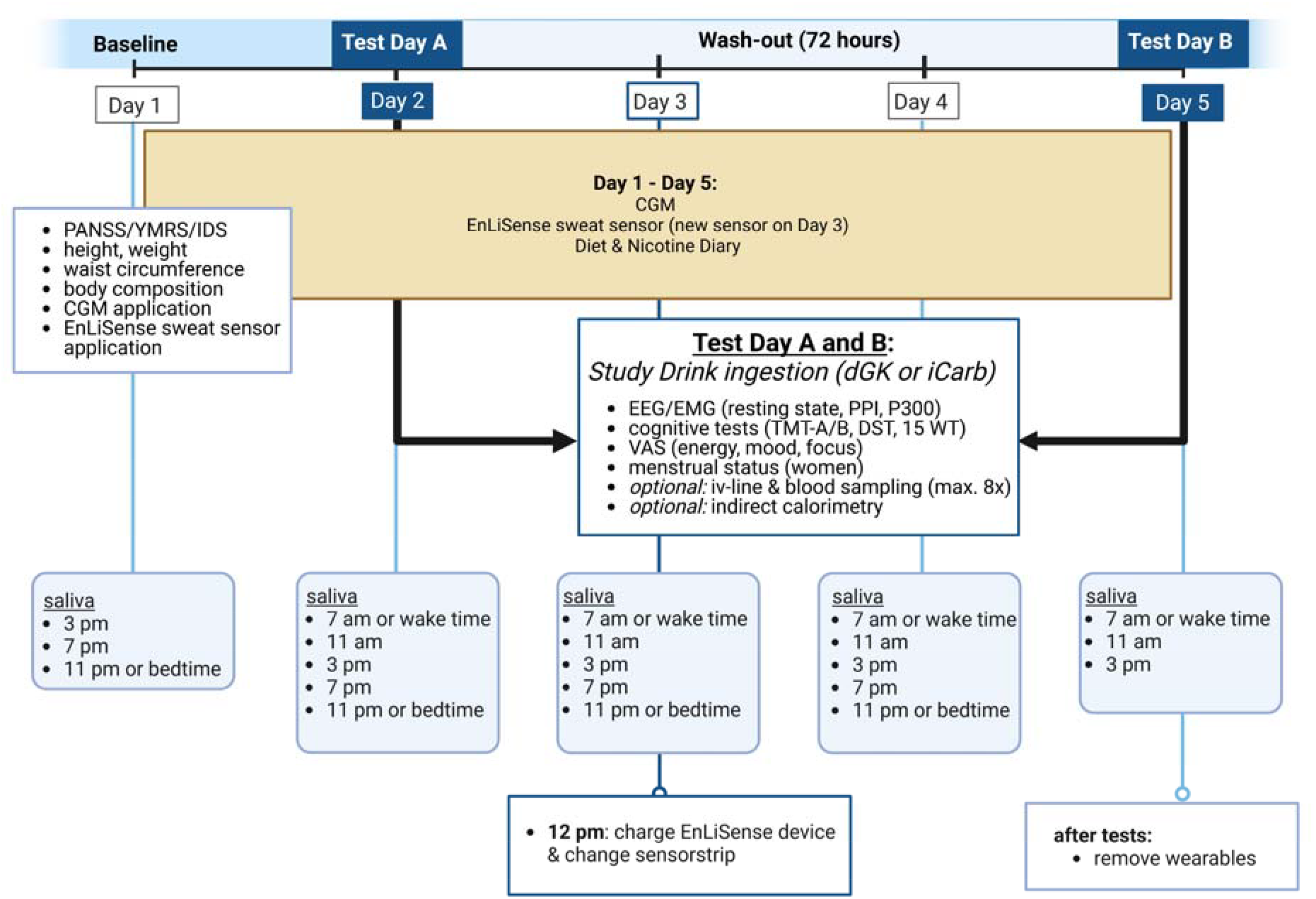
Overview of measurements during study participation.

The study is conducted in an academic hospital setting (Amsterdam UMC, the Netherlands), specifically within the early-episode psychosis ward and acute psychiatry ward of the department of psychiatry. As patients are already hospitalized for the treatment of their psychotic or bipolar episode, they can be closely monitored throughout the trial without requiring additional hospital visits. This minimizes the burden on patients and facilitates their participation in the study.

### 2.4 Study population and eligibility criteria

#### 2.4.1 Study population

The study population consists of patients with recent onset psychosis due to SSD and patients with BD currently having a manic or depressive episode. There are no restrictions concerning gender or ethnicity. All patients receive treatment-as-usual, have significant disease symptoms but are sufficiently recovered to provide informed consent for study participation. The treating medical doctor or psychiatrist evaluates the patient’s mental competence, upon which a researcher obtains informed consent after providing detailed information about the study’s purpose, procedures, potential effects and potential adverse events. The subject information provided to study participants also states that both study drinks are generally considered unpalatable to aid blinding. Patients are given one week to consider participation and receive reimbursement for participation. Participants receive a financial compensation of €200, either via bank transfer or as a gift card.

#### 2.4.2 Inclusion criteria

- The participant has a recent onset psychosis due to SSD, or a manic or depressive episode due to a BD.
- Age 18 years or older.
- Receives treatment-as-usual (including but not limited to antipsychotic and/or mood stabilizing medication).
- The participant is competent to give informed consent.

#### 2.4.3 Exclusion criteria

- Substance use as cause of psychosis or mood episode.
- Substance use (other than nicotine) in the week prior to study onset.
- Intellectual disability (already established at admission to the hospital or clinically assessed).
- Diabetes mellitus type 1 or type 2.
- Metabolic disease impacting ketone metabolism, such as succinyl-CoA:3ketoacid CoA transferase (SCOT) deficiency, multiple acyl-CoA dehydrogenase deficiency (MADD).
- Liver disease.
- Kidney disease.
- Cardiovascular disease.
- The participant is trying to become pregnant, is pregnant or is breastfeeding.

#### 2.4.4 Withdrawal

Subjects can leave the study at any time for any reason if they wish to do so, without any consequences. The clinician or investigator can decide to withdraw a subject from the study for medical reasons. We expect a maximum drop-out percentage of 15% based on previous intervention studies within the same patient groups and setting.

### 2.5 Intervention

### 2.5.1 Investigational product: dGK

The KE (R)-3-hydroxybutyl-(R)-3-hydroxybutyrate (DeltaG® Ketones, dGK) was developed by TDeltaS Ltd. in collaboration with the University of Oxford. dGK is an FDA “generally recognized as safe” (GRAS) approved food product. After ingestion, dGK is hydrolyzed in the small intestine, resulting in the production of BHB and 1,3-butanediol. Both are transported via the portal circulation to the liver, where 1,3-butanediol is metabolized into AcAc and BHB and released into the bloodstream. Currently, dGK has been available on the consumer market as a food product for several years, with extensive safety and toxicology studies conducted prior to its consumer release [62, 63]. No serious adverse events have been reported. Infrequent non-serious side effects include mild gastrointestinal discomfort, dizziness, or headache for a short time [64].

The intervention consists of a nutritional drink containing 50 g of dGK (250 kcal). No correction for body weight is performed, since other factors outweigh body weight in the level of ketosis achieved [65]. The drink is cooled and diluted in water to a total volume of 150 ml to minimize the unpleasant flavor of dGK. After the ingestion of 50 g of dGK the time to reach the maximum peak concentration (Tmax) is between 1-3 hours for BHB, and 1-4 hours for AcAc [63].

#### 2.5.2 Control product: isocaloric carbohydrate drink

A commercially available carbohydrate sports drink, Maurten Drink Mix 160, will serve as the isocaloric control condition. This drink contains maltodextrin and fructose as its primary ingredients. To match the caloric content of the dGK beverage, 63 grams of Maurten Drink Mix 160 (providing 250 kcal) will be dissolved in 150 ml of water, resulting in a 150 ml control drink. Like dGK, Maurten Drink Mix 160 mixed into this volume of water is generally considered to have a very unpalatable flavor and consistency, thus helping with blinding.

### 2.6 Allocation and blinding

Patients are randomized to receive either the dGK or the isocaloric control placebo drink first, using an electronic case report form (eCRF) tool that is accessible via the internet (Castor Electronic Data Capture, castoredc.com). The randomization key is kept confidential, with an independent researcher responsible for preparing and labelling the drinks with codes unknown to patients and test administrators. This researcher is not involved in patient inclusion, test procedures, or data analysis. Both patients and test administrators remain blinded during test sessions. Data analysis is conducted by the study investigators in a blinded manner, with the randomization key revealed only after the analysis is completed, resulting in a triple blind design.

### 2.7 Measures

#### 2.7.1 Baseline characteristics

Several baseline characteristics are collected to form a comprehensive profile of the participants. These include demographic information such as gender, age and ethnicity. Physical measurements are taken: body mass index (BMI), waist circumference and body composition analysis via a bio-impedance scale (Robi S11). Standard clinical laboratory tests, as part of routine care during admission, yield results for complete blood count, electrolytes, C-reactive protein (CRP), and liver, kidney, and thyroid function. Detailed data on medication use, relevant co-morbidities, and other diagnoses are extracted from patient files. For female participants, menstrual phase will be recorded by asking participants about the first day of their previous menstruation, the average duration of their menstrual cycle and any hormonal contraceptives used.

Furthermore, specific clinical interviews are conducted to assess baseline psychiatric symptoms. The Positive and Negative Syndrome Scale (PANSS)[66] is administered to participants with a psychotic episode, the Young Mania Rating Scale (YMRS)[67] for participants with a manic episode, and the Inventory of Depressive Symptomatology (IDS-C)[68] for participants with a bipolar depression.

Participants maintain a dietary diary throughout the study, recording their habitual intake of food and liquids to provide an overview of their dietary habits. Similarly, participants who use nicotine products keep a smoking diary for the duration of the study.

#### 2.7.2 Primary outcome measure

PPI of the acoustic startle reflex reflects the brain’s ability to filter sensory information. When a soft pre-pulse sound precedes a loud, startle-eliciting noise, it inhibits the startle reflex, indicating healthy sensory filtering. A disruption in PPI, where the pre-pulse fails to inhibit the startle reflex, indicates impaired sensory filtering. PPI is considered as one of the of the key neurophysiological abnormalities in SSD and BD, contributing to symptomatic cognitive impairments [69-72]. Combined with the finding that a single dose of exogenous ketones could reverse PPI deficits in an SSD animal model [53], this was reason to select this measure as our primary outcome.

PPI of the startle reflex is assessed using an auditory paradigm with electromyography (EMG) of the orbicularis inferior muscle [73, 74]. The PPI paradigm starts with a 1-minute acclimation period of 70 decibel (dB) white noise (used in the paradigm as background noise), followed by 3 blocks of trials featuring pulse-alone (PA) and pre-pulse-pulse conditions superimposed on the background noise. The pulses consist of white noise sound bursts at 115 dB lasting 20 milliseconds (ms), with intertrial intervals randomly ranging between 10 and 20 seconds. Blocks 1 and 3 measure habituation through 8 PA trials each. Block 2 comprises 50 trials, including 10 PA trials identical to those in blocks 1 and 3. Additionally, block 2 includes four pre-pulse-pulse trials presented 10 times each: pre-pulse sound bursts (76 dB and 85 dB) delivered either 60 ms or 120 ms before the 115 dB pulse. All 50 trials in block 2 are presented in a pseudo-randomized order (no two identical trial types are presented in direct succession). Pulses and pre-pulses are delivered using experiment control software (Presentation®, version 24.1). Participants hear the background noise and stimuli through audiometric insert earphones (3M E-A-RTONE Insert Earphone, ER3A). EEG data are recorded with a calibrated EEG apparatus (EEGO™sports system, ANT Neuro) using EEG caps (Waveguard™original). Participants will wear the EEG cap for the entire duration of the testing, which lasts around 2-3 hours. These caps are equipped with droplead electrodes to measure EMG bipolarly via two electrodes placed below the right eye on the orbicularis inferior muscle. The PPI task will be administered one hour after the intake of the dGK or placebo drink to allow sufficient time for absorption into the bloodstream, and achieve a concentration at near Tmax [75]. The total duration of the PPI task is approximately 20 minutes.

#### 2.7.3 Secondary outcome measures

##### 2.7.3.1 Resting-state EEG

Besides impaired PPI, resting-state EEG alterations are frequently reported in SSD and BD [76-80]. In this study, resting-state EEG recordings are conducted prior to drink ingestion and 30 minutes thereafter. Resting-state EEG will be recorded under first both eyes-open (gaze fixation on visual mark in front of participant) and second eyes-closed conditions, with each condition lasting 5 minutes. Participants are instructed to remain seated, maintain an upright posture, relax, and minimize movement during the recording. We investigate power spectral differences, specifically increased slow oscillatory power (delta/theta) in the frontal regions.

##### 2.7.3.2 P3B Event-Related Potential

The P3B amplitude is a specific type of event-related potential (ERP), an indicator of cognitive processes such as attention, working memory and decision-making. P3B amplitude is considered a state and trait marker in SSD [81]. Both SSD and BD are associated with reduced P3B amplitude and delayed latency [76, 82], yet the potential effects of ketone bodies on P3B amplitude remain unexplored. P3B amplitude is often measured by providing a random visual or auditory oddball paradigm where participants respond to infrequent stimuli in a sequence of a frequently occurring (standard) stimulus [81]. During our P3B task, 160 stimuli are presented in random order, with participants instructed to press a button upon detection of the ‘oddball’ target (32 out of the 160 stimuli). The task is preceded by 10 practice trials. The standard (nontarget) stimulus consist of a 500 Hz pure tone, while the ‘oddball’ target stimulus is 700 Hz, each with a duration of 100 ms. Both stimuli are presented at an intensity of 80 dB, with a random interstimulus interval ranging from 1.2 to 1.5 seconds. Throughout the P3B task, brain activity is recorded using the same EEG equipment as mentioned above to capture neural responses (Pz channel), following the presentation of these stimuli.

##### 2.7.3.3 Cognitive Outcome Measures

Cognitive impairments in SSD and BD have been extensively studied, highlighting deficits in cognitive domains such as executive functioning and memory [83]. After the above-mentioned neuroelectrophysiological measurements, the effects of the drink on cognitive functioning are assessed using three validated neuropsychological tests, reflecting various aspects of cognitive functioning. The Trail-Making Tests (TMT-A and TMT-B, 2007, Dutch translation; paper versions) measure visual-motor speed, attention, and cognitive flexibility, with TMT-B also reflecting executive functions such as task-shifting [84]. The Digit Span Test (DST), as part of the Wechsler Adult Intelligence Scale (4^th^Edition), evaluates auditory working memory capacity, both in terms of forward and backward digit recall, as well as cognitive control [85]. The 15-Word Test (15 WT; a Dutch adaptation of the Rey Auditory Verbal Learning Test) assesses short- and long-term verbal memory through immediate recall, delayed recall, and recognition of previously presented words, providing a comprehensive measure of memory processes [86]. Parallel tests are used to minimize learning effects, which are otherwise compensated by the AB/BA crossover design of the study.

##### 2.7.3.4 Patient-experienced and reported outcomes

Participants are asked to rate their energy level (from very exhausted to very energetic), mood (from very depressed to very happy), and ability to focus (from very poor to very well) using Visual Analog Scales (VAS) at two timepoints: before and 2 hours after ingestion of the nutritional drink, on both test days.

##### 2.7.3.5 Immune, oxidative stress and metabolic blood parameters

Participants may opt out of blood sampling. For those who consent, a venous cannula is inserted into a forearm vein at the start of each session to facilitate sequential blood draws without repeated venipunctures. Blood is collected for serum and plasma isolation at the following time points: before drink ingestion (baseline, T0), at 20-minute intervals during the first hour after ingestion (T1-T3), and at 30-minute intervals thereafter (T4-T7), with a maximum of 8 blood draws per session. At all timepoints, a separate blood sample is taken and put on ice for ketone body determination, preventing conversion into and evaporation of acetone. In addition, a Tempus Blood RNA tube is collected at 1 hour (T3) after drink ingestion, for gene expression analysis using the metabolic and immune nCounter® gene expression panels from Nanostring (Seattle). All samples arestored at -80°C until further analysis.

Immune, oxidative stress and metabolic markers are determined in serum, plasma and deproteinized blood. Inflammatory markers include at least interleukin-6 (IL-6), interleukin-10 (IL-10), high-sensitivity C-reactive protein (hsCRP), and soluble intercellular adhesion molecule-1 (sICAM1) [13, 20, 87-89]. Oxidative stress markers include glutathione S-transferase (GST) and superoxide dismutase (SOD), key enzymes in ROS regulation [90, 91]. Metabolic markers at least include glucose, ketone bodies (BHB and AcAc) and acylcarnitine profile [92, 93].

##### 2.7.3.6 EnLiSense sweat sensor and continuous glucose measurement

Participants will wear a non-invasive sweat sensor (EnLiSense LLC, Dallas) throughout the 5-day study period, enabling near-continuous multiplex monitoring of biomarkers in sweat. This study focuses on markers linked to circadian rhythms and inflammation. Studying circadian rhythms is challenging due to the invasive nature of frequent melatonin and cortisol sampling, the primary regulators of circadian rhythms [94]. The ability to continuously measure cortisol and melatonin levels could provide deeper insights into the circadian rhythms of patients with SSD and BD [95]. Moreover, evidence from *in vivo* studies suggests that the KD may influence circadian rhythms [96], and it has been hypothesized that improvements in the sleep-wake cycle could mediate the beneficial effects of ketone bodies in SSD and BD [97]. Furthermore, IL-6 and tumor necrosis factor-α (TNF-α) will be measured, as these pro-inflammatory cytokines are often be elevated in patients with psychosis [13, 89]. In an acute inflammation mouse model, KE prevents increased expression in the brain of pro-inflammatory cytokines involved in SSD (IL6, TNFA, IL1B) [98], suggesting KE could normalize levels of these cytokines in SSD/BD.Therefore, assessing whether these markers change following ketone drink consumption could provide valuable insights into its potential effects on inflammation.

Melatonin, cortisol, IL-6 and TNF-α are determined in saliva samples as a validation for the sweat sensor data (Ella™ Automated Immunoassay System, Bio-Techne®). Saliva samples are collected at fixed time points daily: 7.00 a.m. (or when the participant wakes up), 11.00 a.m., 3.00 p.m., 7.00 p.m. and 11 p.m. (or before bedtime). Saliva is collected in Eppendorf tubes® by passive drool method (BioSaliva collection aid). Saliva samples are stored at -80°C until further analysis.

Additionally, participants wear a continuous glucose monitor (CGM, Abbott Freestyle Libre 2) for the entire five-day duration of the study to gain further insight into their glucose metabolism [89, 99].

##### 2.7.3.7 Indirect Calorimetry

Information on substrate utilization and energy metabolism isobtained through indirect calorimetry. Indirect calorimetry is an optional test procedure for study participants. This method involves measuring oxygen consumption and CO_2_production using a ventilated hood system (Q-NRG, Cosmed), which allows for the automatic calculation of resting energy expenditure (REE) and the respiratory quotient (RQ). REE reflects the energy metabolism of the participant at rest. The RQ (the ratio of CO_2_exhaled to O_2_consumed) provides insights into whole-body substrate oxidation, including glucose, fat, and protein. Participants are required to remain still, but not sleep, beneath the ventilated hood system for approximately 15 minutes, after which the REE is calculated. Substrate-oxidation is calculated as previously described [100].

### 2.8 Data collection and management

Researchers involved in the study have received expert training in conducting PANSS, YMRS, and IDS-C interviews, performing electrophysiological measurements, administering cognitive tests, applying sensors, collecting blood and saliva samples, .

Each participant is assigned a unique code for data storage, handling, and analysis, which cannot be directly traced back to the individual. Access to the identification key is restricted to authorized personnel. Data collection forms are completed on paper and subsequently entered into the eCRF (Castor EDC). To ensure data quality, assessors are thoroughly trained in Good Clinical Practice (GCP). Paper forms, including signed informed consent documents, are securely stored in a locked filing cabinet. Digital data are saved in a secured folder with restricted access for research personnel. Data will be retained for 15 years following the study’s conclusion. Anonymous, selected material and data is shared after study conclusion with collaborators at the University of Texas (Department of Bioengineering, Dallas, USA) and the University of Alberta (Department of Pediatric Cardiology, Edmonton, Canada) through Material and Data Transfer Agreements. Anonymous EnLiSense sweat sensor data is shared with EnLiSense LLC through a Data Transfer Agreement. All shared data and samples remain coded to protect participant confidentiality. An independent monitor performs quality assurance.

### 2.9 Data Analysis

#### 2.9.1 Sample size

This study is the first to investigate the effects of KE in SSD and BD patients, and is designed as a pilot and feasibility study, precluding a formal sample size calculation based on pre-existing data. Due to the lack of consensus on sample size calculation in pilot/feasibility studies, we have relied on existing literature as a basis for our decision. Following Julious’ recommendations [101], we have chosen a sample size of 12 subjects per group. Therefore, a total of 24 patients with either SSD (n = 12) or BD (n = 12) will be included in the study. If a participant drops out, efforts will be made to include a replacement.

### 2.9.2 Statistical analysis

Comparisons regarding all outcome measures within patients (dGK condition versus control drink condition) are performed using paired-samples Student’s t-tests for normally distributed data (or Wilcoxon signed-rank test if parametric assumptions are not met). Group-level effects will be analyzed using a linear mixed effects model, with adjustment for confounding factors. A significance level of ≤0.05 Given the exploratory nature of this pilot study, no imputation analysis will be conducted in the case of missing data (except for missing EEG channels) and no correction for multiple testing will be applied.

### 2.10 Data monitoring

#### 2.10.1 Patient safety

This study utilizes well-established nutritional drinks with no known harmful side effects. Patients are clinically admitted to the department of psychiatry and are closely monitored by both the investigators and the clinical staff responsible for their treatment. In the highly unlikely event of an unexpected adverse effect, the procedures outlined in the study protocol will be followed. In accordance with Section 10, Subsection 4, of the WMO, the sponsor will suspend the study if there are sufficient grounds to believe that continuing the study may endanger the health or safety of the subjects.

#### 2.10.2 Public disclosure and publication policy

Results from the study will be presented at patient information events (e.g., hosted by patient organization *Anoiksis*, members of which contributed to the design of this study). Additionally, manuscripts about the obtained results (regardless of whether the results are positive, negative or neutral) will be submitted for publication in peer reviewed international journals.

## 3. Discussion

Our current study investigates the effects of KE compared to an isocaloric carbohydrate control drink in patients with SSD and BD. To the best of our knowledge, this is the first study to examine the impact of KE in this patient population. While a few uncontrolled studies suggest beneficial effects of a KD on SSD and BD, dietary interventions can be challenging to study in a controlled setting [97, 102-106]. In the context of SSD or BD, adhering to a ketogenic diet poses an additional challenge, as core symptoms associated with cognitive deficits, negative symptoms like loss of motivation and disorganization may decrease compliance. The challenging nature of a KD can also lead to selection bias, favoring patients with a lower disease burden and strong social support, thereby limiting generalizability. Importantly, during the acute disease phase, adherence to a strict diet is not feasible. Moreover, it takes several days to weeks to reach sufficient levels of ketosis after starting the KD, rendering this intervention unsuitable to achieve fast ketosis, and thus unsuitable as a treatment strategy in the acute disease phase. Finally, psychiatric medications can inhibit ketogenesis, making it hard for patients to reach ketosis on a KD [107]. KE offers an interesting alternative, yielding acute, titratable ketosis. Studies using KE can therefore facilitate a more rigorous investigation of the effects of ketone bodies in SSD/BD. Additionally, if effective, they could provide a viable therapeutic strategy for SSD/BD patients who struggle with dietary adherence, thereby making ketosis-based interventions more accessible to a diverse patient population.

Our study has several strengths. We consider the broad range of outcomes a strength, since it may inform future trials. Ketone bodies are hypothesized to improve symptoms of SSD/BD through multiple mechanisms. In this pilot study, we focus primarily on their effects on frequently observed neuro-electrophysiological abnormalities in SSD and BD (i.e., impaired PPI [69, 70], resting-state EEG changes [76-80], and reduced P3B amplitude [76, 81, 82]). By assessing these neuro-electrophysiological outcomes, alongside perceived mood, energy levels, cognitive function and metabolic, oxidative stress and immune parameters, our current pilot study seeks to understand how ketones may influence brain function in SSD and BD patients. Ketone bodies may also improve symptoms of SSD/ BD through less explored mechanisms, such as the circadian rhythm. While a single administration of KE may not impact the circadian rhythm, continuously monitoring cortisol and melatonin levels in this study could provide deeper insights into the circadian rhythms of patients with SSD and BD.

Importantly, previous studies often lack consistent measurement of blood ketone levels. In this study, we include sequential measurements of blood ketone levels, providing detailed insights into the dynamics of BHB and after KE ingestion. We expect that the induced hyperketonemia may lead to effects similar to those observed during endogenous ketosis – when ketone bodies are produced by the body itself – based on previous research demonstrating a metabolic competition between ketone bodies and glucose, favoring ketone utilization even in the context of an unchanged diet [27, 58].

The randomized AB/BA cross-over design of this study is expected to strengthen trial outcomes. In a cross-over trial, participants function as their own controls, enhancing both the interpretability of results and the statistical power despite the limited sample size. Moreover, this design reduces interpatient variability in the comparison between groups and the effect of covariates such as participants’ demographic characteristics. Randomization to AB/BA can compensate for test-retest bias and potential carryover effects. By choosing electrophysiological outcome measures (relatively insensitive to test/retest effects), we further reduce bias in this crossover design.

A limitation of the current pilot study is the use of a single ingestion of dGK, instead of a longer period of supplementation. This limits the outcome measures to parameters with an expected acute response to ketosis. For instance, our current study cannot answer if negative symptoms would improve with KE. We aim to investigate this important question in future studies.

A known limitation of pilot studies investigating novel concepts is the lack of prior data, which prevents the ability to perform an informed power analysis. This, in turn, can weaken the resulting recommendations or conclusions. However, this gap in knowledge is precisely why our current study is necessary. It will provide the data needed to inform a subsequent randomized controlled trial, including a comprehensive power analysis. Another potential limitation is that do not assess whether the participants have a baseline deficit in PPI. It is known that not all SSD and BD patients exhibit PPI deficits, which could limit the outcomes of our intervention. However, in this study, we will measure the percentage change in PPI after dGK ingestion compared to an isocaloric carbohydrate control drink, allowing each participant to serve as their own control thereby mitigating the effects of this limitation.

There are several biases that need to be considered in this study. The Department of Psychiatry at Amsterdam UMC functions as a regional hospital for acute psychiatric episodes, such as manic episodes or psychosis. The Early-Episode Psychosis ward primarily admits patients who agree to their treatment and are willing to discontinue street drug use. This may introduce selection bias, as it represents only a subset of SSD/BD patients. Conversely, including a more heterogeneous group could introduce additional confounders and affect the results. In addition, the substantial number of outcome measures in a moderately affected SSD/BD patient population presents a challenge, as it requires several (up to four) hours of intensive testing per test day. This could potentially lead to attrition biases, with more vulnerable patients dropping out of the study. However, we currently successfully finished data acquisition for 7 patients with SSD/BD in our study, without drop-out.

In conclusion, our current study is a first step in exploring the potential benefits of KE to improve symptoms in SSD and BD. By investigating the effects on neuro-electrophysiological outcomes, cognitive performance, metabolic, immune, oxidative stress, and circadian rhythm parameters, we aim to deepen our understanding of how ketones influence fundamental brain function related to behavioral and symptomatic manifestations in these patient groups. Our current study is a pilot aimed at informing future larger-scale research with a longer period of supplementation and longer follow-up evaluating the potential of KE to improve symptoms in SSD and BD. If proven effective, KE could offer a novel therapeutic approach to modify pathophysiological mechanisms and thereby reduce functional impairments and alleviate the burden of these disorders.

## Data Availability

On completion of the study data anonymized data can be made available at the discretion of the authors, upon written request.

## 4. Author Contributions

Conceptualization KH, NvB, LH, MS

Methodology KH, YY, BO, DS, TZ, MT, SP, SM, MP, BR, RH, RK

Software RMO, DD, DS, BO, MP

Validation KH, DD, YY, RMO, DS, BO, MP

Formal analysis AK, DD

Investigation DD, RMO

Resources DS, BO, LH, SP, SM, MS, JD

Data Curation DD, DS, BO, RMO

Writing - Original Draft DD

Writing - Review & Editing KH, ED, NvB, LH, MT, BO, DS, MS, AK

Visualization KH, DD

Supervision KH, YY, MP, NB, LH

Project administration KH, MP, YY

Funding acquisition KH, NB, LH

## 7. Data availability statement

